# Validation and Psychometric Properties of a Depression Scale for People with Tuberculosis in Lima, Peru

**DOI:** 10.1101/2025.08.05.25332532

**Authors:** Paulo Ruiz-Grosso, Johann Vega-Dienstmaier, Cesar Ugarte-Gil

**Author notes:** Validation of a Depression Scale in TB Patients. <u>Corresponding author:</u> Name: Paulo Ruiz-Grosso, <u>Address:</u> Eloy Espinoza Saldaña 709, San Martín de Porres 15102, Lima, Peru <u>Full contact details:</u>.

## Abstract

**Background:** Depression is a known risk factor for poor outcomes during tuberculosis (TB) treatment. Assessing depressive symptoms in TB patients is challenging due to overlap with symptoms of the disease and side effects of treatment, such as fatigue, sleep disturbance, and appetite changes. This study aimed to develop and validate a screening scale for major depressive episodes in patients undergoing TB treatment.

**Methods:** A cross-sectional validation study was conducted using 58 items drawn from previously validated Spanish-language depression scales. These items were administered to adults receiving TB treatment in Lima, Peru. Diagnosis of a major depressive episode was determined through a SCID-5 interview conducted by a psychiatrist. Items were selected sequentially based on individual area under the receiver operating characteristic curve (auROC), with the final selection made when no further gain in discrimination was observed. Internal consistency and diagnostic performance were assessed.

**Results:** Among 163 participants, an 8-item scale achieved an auROC of 95.9% (95% CI: 92– 99.9%), sensitivity of 87.2%, and specificity of 92.2% at a cutoff score ≥11. Internal consistency was good (Cronbach’s α and McDonald’s ω = 0.804). Items on anhedonia, energy, sleep, and appetite were excluded.

**Conclusion:** The validated 8-item scale showed strong discriminative accuracy and internal consistency for detecting major depressive episodes in TB patients.

## Introduction

Tuberculosis (TB) is a global public health issue, especially in low- and middle-income countries ^1^. Drug-resistant strains contribute significantly to its burden, with 8.33% and 1.47% of tested patients in 2021 classified as MDR-TB and pre-XDR/XDR-TB, respectively ^1^. Previous treatment failure or loss to follow-up are key factors linked to MDR-TB^2^, and these are often associated with mental health disorders, particularly substance use and depression ^3–6^.

The prevalence of depressive disorders varies substantially across populations and settings, ranging from 16% at any point during treatment to 38–54% at the start of TB treatment, and decreasing to 7–33% by treatment completion in drug-sensitive and MDR-TB patients, respectively. The presence of depressive symptoms consistent with a major depressive episode (MDE) at treatment onset has also been associated with adverse TB outcomes (HR = 3.54; 95% CI: 1.43–8.75), a finding consistently reported in the literature, particularly for loss to follow-up (OR = 8.70; 95% CI: 6.50–11.64) ^7–13^.

Although locally and internationally validated scales are frequently used in clinical and epidemiological research, most include items assessing somatic and cognitive symptoms (e.g., concentration, memory) that may be influenced by TB itself. For instance, items related to appetite, daily functioning (CES-D 20-item scale, items 2 and 7), sleep disturbances, or fatigue (PHQ-9, items 3 and 4) may reflect TB symptoms rather than depression ^14–17^. This challenge mirrors the rationale behind the development of the Edinburgh Postnatal Depression Scale, whose authors noted that existing scales overestimated depression due to physiological changes in the postpartum period.

Similarly, items related to worthlessness or hopelessness may be influenced by the economic disruption often accompanying TB diagnosis and treatment ^18,19^. Additionally, anti-TB medications can cause appetite and sleep changes, or even depressive symptoms ^20^. These symptom overlaps may lead to overestimating MDE prevalence, potentially misclassifying individuals as depressed and prompting unnecessary mental health interventions, further straining already burdened health systems. To address this issue, the present study aims to identify and select items from a pool of existing depression scale items based on their discriminative performance in a population undergoing TB treatment in Lima, Peru.

## Methods

### Study design and setting

This study was designed as a cross-sectional validation study. Participants were individuals diagnosed with pulmonary tuberculosis confirmed by smear microscopy or culture, and undergoing treatment in accordance with the guidelines of the National Tuberculosis Control Program (PCT). Patients were recruited from health centers operated by the Ministry of Health of Peru. For this study, six primary care health centers and two secondary-level maternal and child health centers located in the jurisdiction of the Lima Norte Health Network Directorate (DIRIS Lima Norte) in Lima, Peru, were selected.

### Study population and sampling strategy

The study population comprised individuals from lower-middle to very low socioeconomic backgrounds, many of whom are either internal migrants or descendants of migrants from northern Peru.

All individuals meeting the following inclusion criteria were invited: (1) pulmonary TB diagnosis confirmed by smear or culture, (2) undergoing treatment for pulmonary TB, and (3) aged ≥18 years as per their National Identity Document (DNI). Exclusion criteria included: (1) diagnosis of extrapulmonary TB, or (2) any condition, as determined by the treating physician, that could impair proper data collection or independent completion of self-administered instruments. A consecutive sampling method was used, recruiting eligible participants successively until the required sample size was reached. TB Control Program (PCT) staff at each of the eight participating health centers notified the study team of potential participants, initiating the recruitment process.

### Exposures, interventions, outcomes

#### Index test

To develop the initial test scale, three expert psychiatrists were asked to propose freely available instruments for item selection. The selected tools included the CES-D, the Depressive Psychopathology Scale, the Zung Depression Scale, the Edinburgh Postnatal Depression Scale, the Hamilton Depression Inventory, and the PHQ-9.

Overlapping items were consolidated to avoid redundancy, and additional items were proposed by experts to enhance the scale’s ability to discriminate depressive episodes in the clinical context of tuberculosis care. This process resulted in a 58-item self-administered questionnaire.

#### Comparison test

The module for major depressive episode of the Structured Clinical Interview for DSM-5 (SCID-5) was used as the gold standard. This instrument provides a structured guide for diagnostic interviews conducted by trained health professionals. The interviews were conducted by PRG, who was the responsible for the patients’ routine care.

### Data collection and measurement procedures

Screening was conducted by trained study staff using a card with inclusion/exclusion criteria. Eligible individuals were invited to speak with the team after a brief explanation. If interested, an appointment was scheduled for the same day. During the visit, study aims and procedures were explained, and informed consent was obtained. Participants first completed self-administered instruments in a private setting, with staff available for support. Subsequently, a psychiatrist (PRG) conducted a structured interview for depression and other diagnoses. All procedures were completed in a single session (∼1 hour). Participants were thanked and provided with contact information for further inquiries.

### Ethical considerations

The study protocol was approved by the Institutional Ethics Committee of Universidad Peruana Cayetano Heredia. All participants provided written informed consent.

### Statistical analysis

Sociodemographic and clinical variables were described using proportions and measures of central tendency and dispersion. To identify the most discriminant items from the initial pool of 58, we applied a method in which each item was individually evaluated by estimating its area under the receiver operating characteristic curve (auROC) for the detection of Major Depressive Episode (MDE), as defined by the SCID-5. The item with the highest auROC was selected as the first item in the scale. This process was repeated iteratively, with each subsequent step adding the item that produced the greatest incremental increase in auROC. The procedure continued until no further improvement in auROC was observed. This selection method, based on auROC optimization, will hereafter be referred to as the S-ROC method.

Exploratory analyses included the estimation of the Cronbach’s alpha and McDonald’s omega to evaluate internal consistency. Exploratory factor analysis and network analysis were conducted to examine the underlying structure of the scales, and item response theory (IRT) was applied to evaluate item discrimination and difficulty.

Statistical analyses were performed in Stata v19.0, except for network analyses, conducted in xxx xxx. Details are in Supplementary Material.

Based on a 5% significance level, 80% power, 30% disease prevalence, and sensitivity increasing from 0.60 to 0.80, a sample of 45 cases and 150 non-cases was estimated using PASS 2021, following *Bujang et al.* ^21^

## RESULTS

Data from 163 participants across 14 health centers showed that 38.3% were treated for drug-sensitive TB, 46.3% for MDR-TB, and 2.68% for XDR-TB. Median age was 31 years, 46.3% were female, and median education was 11 years (IQR = 9.5–14). Most were single (49.7%), 29% were born outside Lima, and had lived in the city for a median of 20 years. The most common occupations were student (18.5%), merchant (16.1%), and skilled worker (15.4%). Mental health treatment history was reported by 44.9%, and 23.2% had a relative in treatment (Table 1).

**Table 1.** Sample description (n=158)

| **Variable** | **Category** | **N (%)** |
| --- | --- | --- |
| **Sex** | Female | 73 (46.2) |
|  | Male | 85 (53.8) |
| **Age \*** |  | 31 (23-45) |
| **Marriage status** | Single | 76 (49.7) |
|  | Married or living with a partner | 53 (36.0) |
|  | Divorced or widowed | 21 (14.3) |
| **Years of study \*** |  | 11 (9.5-14) |
| **Job** | None, recycler | 13 (10.0) |
|  | Professional job | 4 (3.1) |
|  | Construction/Contractor | 8 (6.1) |
|  | Commerce | 21 (16.1) |
|  | Independent work | 20 (15.4) |
|  | Transportation | 13 (10.0) |
|  | Professional | 15 (11.5) |
|  | Student | 24 (18.5) |
|  | Homemaker, caregiver, or cleaner | 12 (9.2) |
| **Place of birth** | Lima | 93 (71.0) |
|  | Coast | 13 (9.9) |
|  | Andean | 16 (12.2) |
|  | Jungle | 6 (4.6) |
|  | Abroad | 3 (2.3) |
| **Years living in Lima \*** |  | 20 (9.5-30.5) |
| **Previous mental health treatment** |  | 66 (44.9) |
| **Previous mental health treatment of a family member** |  | 33 (23.2) |
| **Treatment regimen** | Sensitive | 57 (38.26) |
|  | MDR | 69 (46.31) |
|  | XDR | 4 (2.68) |
|  | RAFA | 5 (3.36) |
|  | Other | 14 (9.4) |

\*median(IQR)

In the SCID-5 interview, 30.6% met criteria for Major Depressive Episode (MDE) as defined by at least two major symptoms and five total symptoms. Additionally, 20.9% received a clinical diagnosis of MDE and 44.7% had any clinically significant depressive disorder. The most commonly reported symptoms were fatigue/loss of energy (54.1%), cognitive difficulties (40.9%), and sleep disturbances (38%) (Table 2).

**Table 2.**
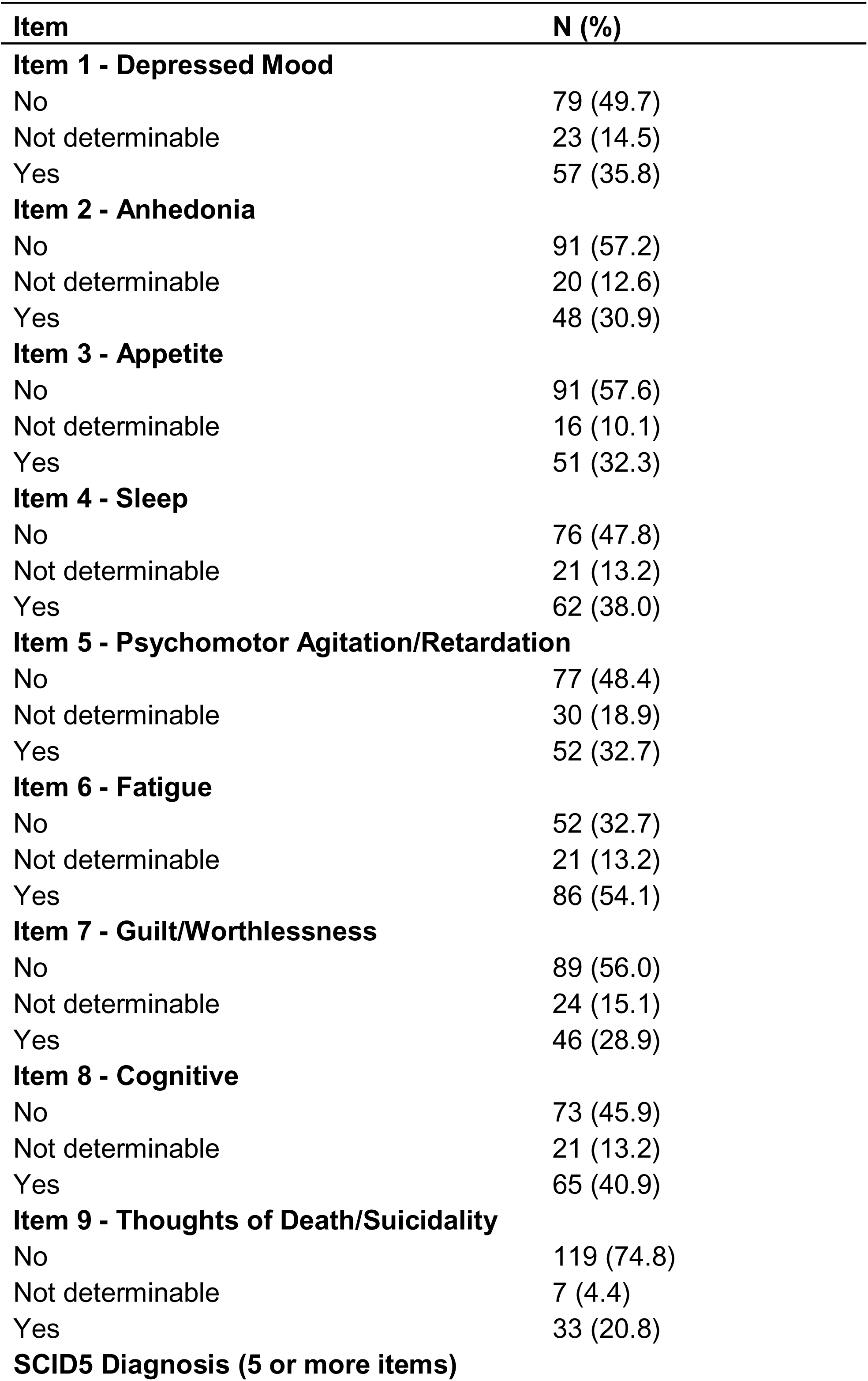
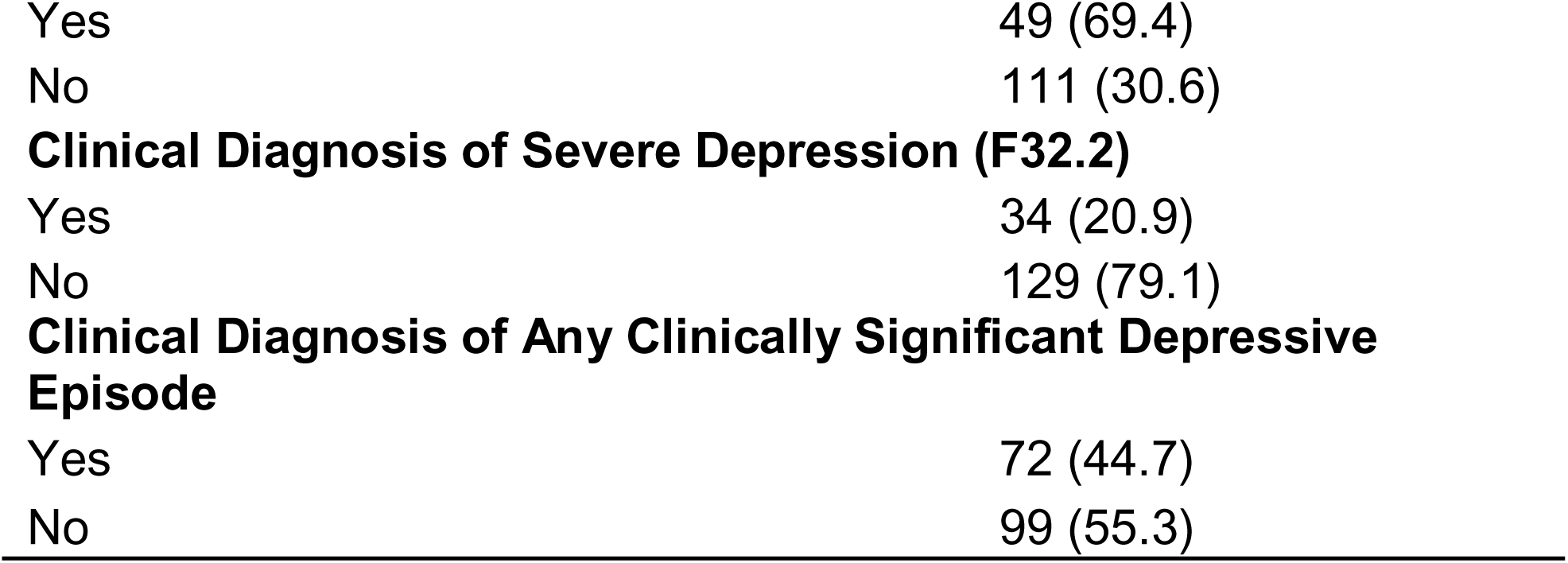
Depressive symptoms and disorders in patients receiving treatment for TB according to the SCID5.

### Main and exploratory analyses

Using the S-ROC method in the initial pool of 58 items, the resulting 8-item scale included: i2 “felt depressed,” i48 “concentration,” i24 “better off dead,” i47 “difficult thinking,” i31 “restless,” i33 “worried,” i21 “lost patience,” and i46 “happy with good news.” This scale showed an auROC of 95.9% (95% CI: 92.0%–99.9%), with an accuracy of 90.67% at a cutoff score of ≥11, yielding a sensitivity of 87.23%, specificity of 92.23%, and a Youden’s J of 0.795. Its auROC was significantly different than that of the original 58-item pool (p<0.001). See Table 3 and Figure 1

**Figure 1.**
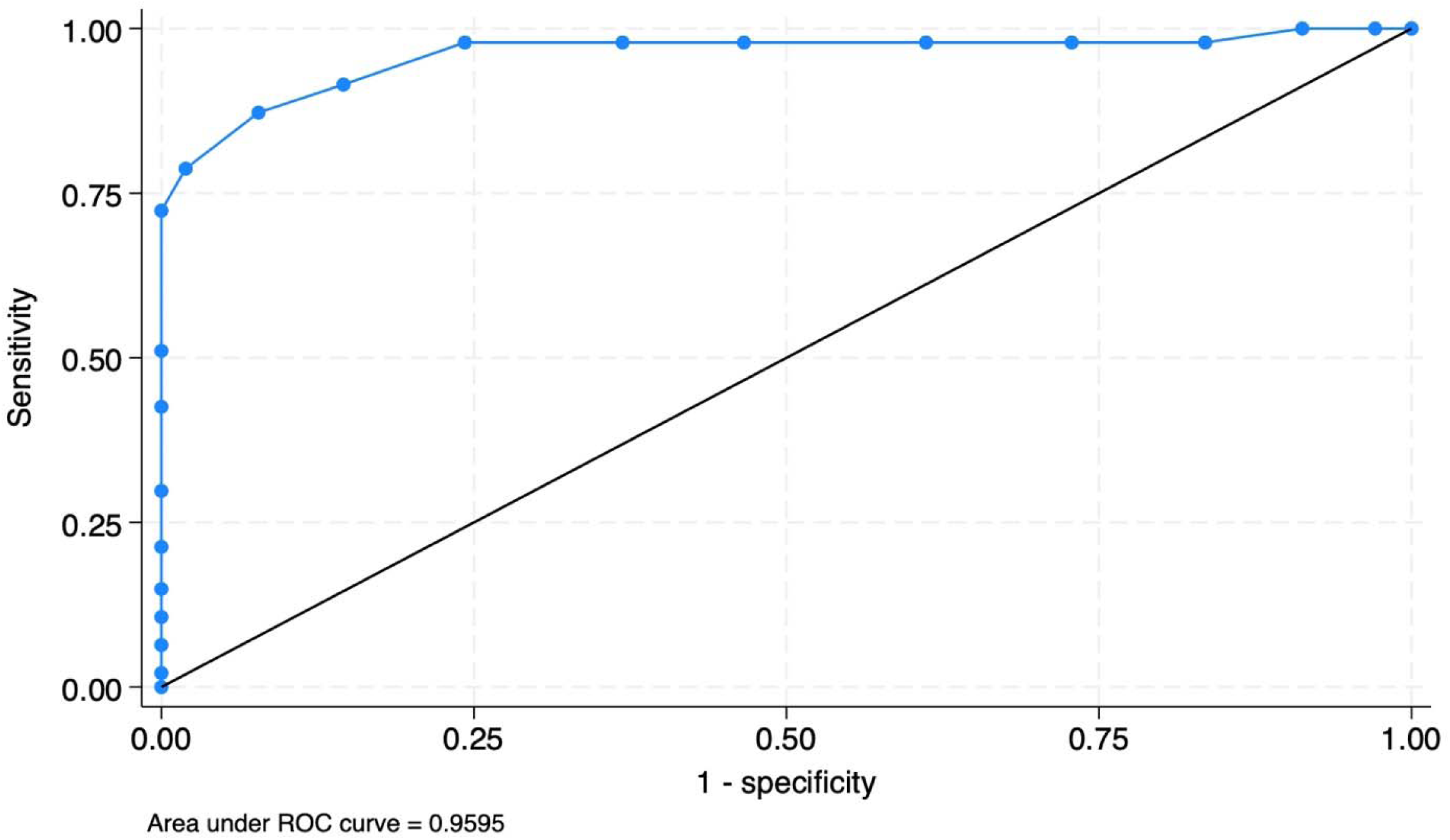
ROC Curve of 8 item scale.

**Table 3.**
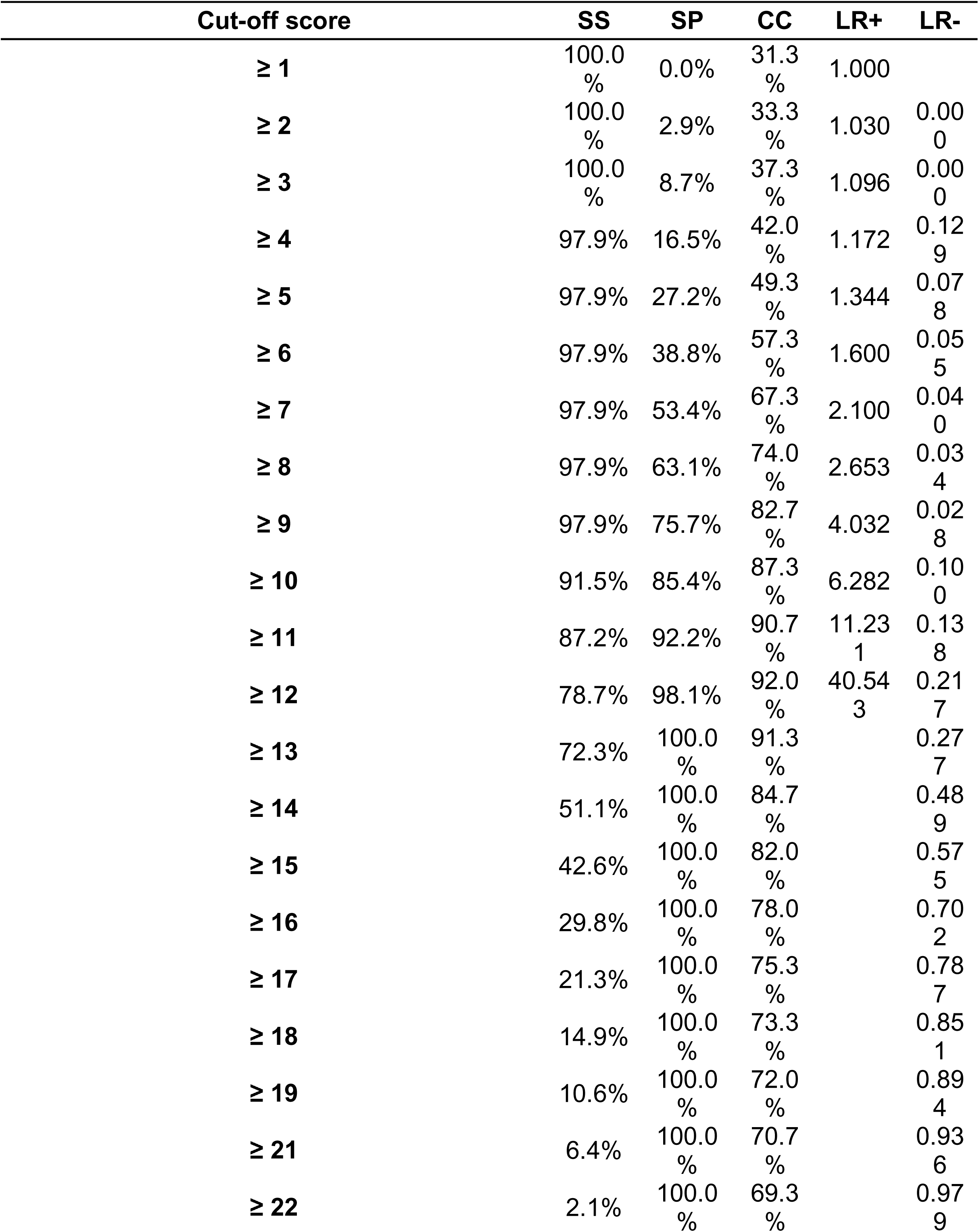
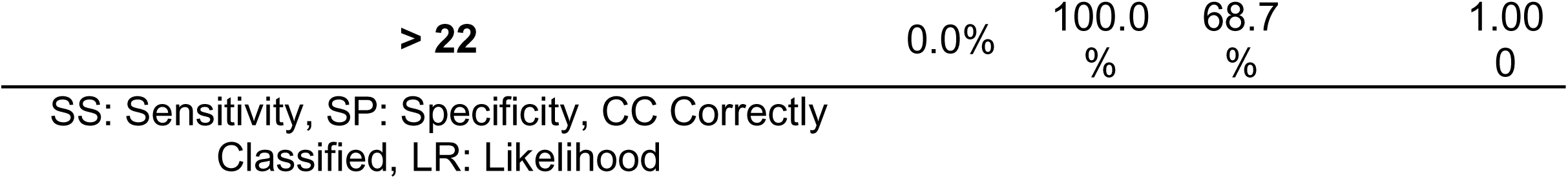
Diagnostic performance of total score cutoffs for detecting MDE.

Internal consistency was acceptable (Cronbach’s α and McDonald’s ω = 0.804). The EFA showed evidence of adequate psychometric properties, including lack of multicollinearity (Bartlett’s test, p < 0.05) and adequate sampling adequacy (KMO = 0.819). Two items 48 and 46 loaded on a “positive mood” factor, while the remaining items loaded on a factor related to “anxious and depressive symptoms.” Only item 21 had a factor loading below 0.4. Item 2 (“felt depressed”) contributed most to internal consistency (Δα = 0.045), had the highest expected influence in network analysis (EI = 1.867), and the highest discrimination parameter (a = 2.45) in item response theory analysis. See Table 4 and Figure S1.

**Table 4.**
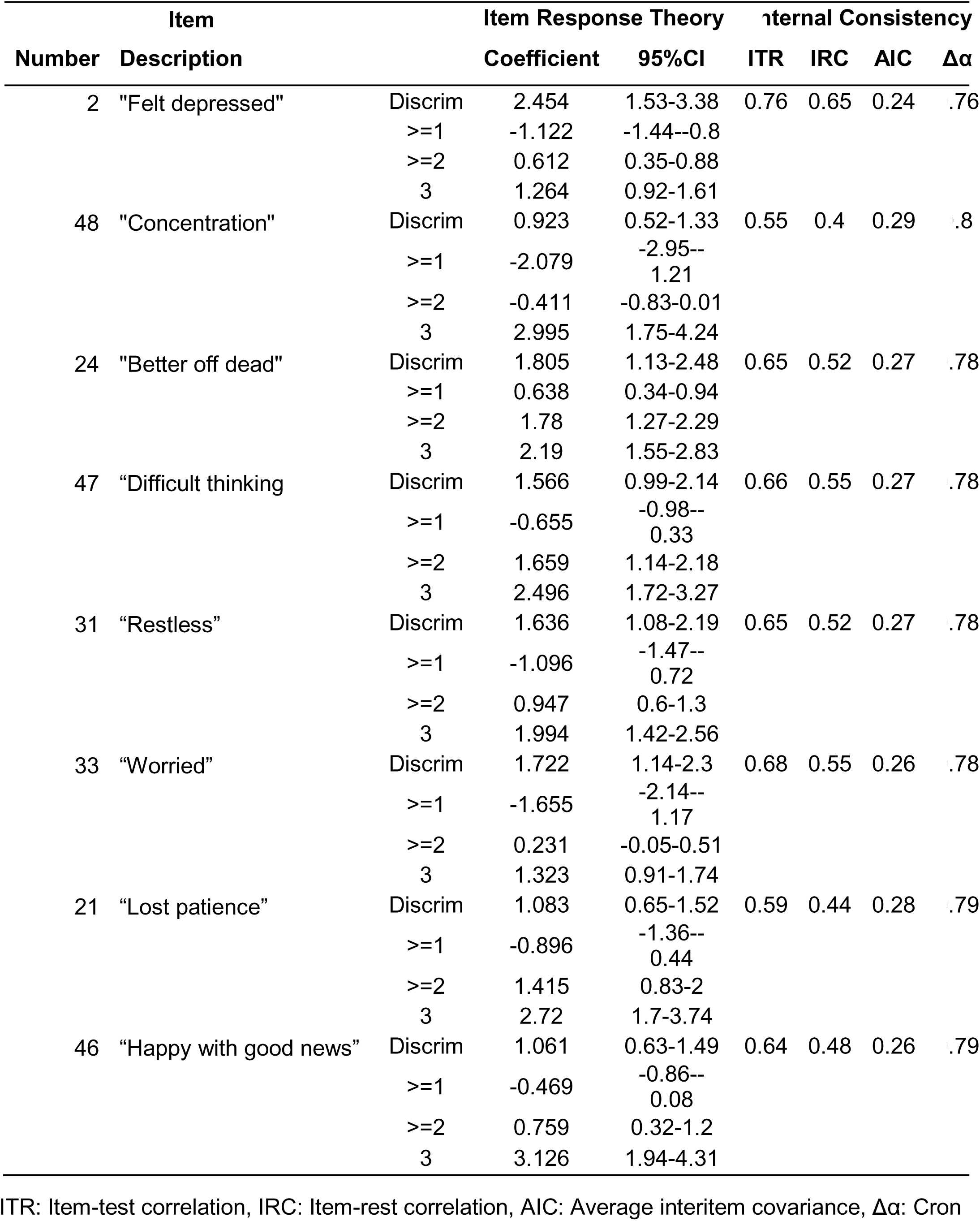
Item response theory, internal consistency and network analyses estimates.

## DISCUSSION

Our results suggest that, in a population of individuals undergoing TB treatment, a scale composed of 8 items—derived through an iterative selection process from a pool of items commonly found in established depression scales—exhibits good psychometric properties for the detection of MDE, using the SCID-5 as the gold standard. The optimized 8-item scale effectively captures core depressive and anxious symptoms and may reflect an underlying structure that distinguishes between negative and positive affect. Its strong psychometric performance—demonstrated by high internal consistency, a coherent factor structure, and strong discriminative capacity—supports its potential as a brief and practical screening tool for MDE in TB-affected populations.

The consistent performance of item 2 (“felt depressed”) highlights its central role in detecting clinically significant depression. It directly overlaps with the first item of the PHQ-2 and conceptually aligns with restlessness and sadness in the K6, reflected in our items 2 and 31 ^22^. Among locally validated scales, only item 46 (mood reactivity) from the 4-item Depressive Psychopathology Scale overlapped, though both scales addressed depressed mood (sadness vs. “felt depressed”) as a core symptom ^23^. The 5-item CES-D included depressed mood, loneliness, and feelings of failure, with only depressed mood (item 2) aligning with our scale ^14^. These findings suggest that depressive mood is the most consistent anchor across short scales, while other core DSM-5 and ICD-10 symptoms, such as anhedonia and fatigue, are often absent.

When further comparing the items selected for our scale with those of the PHQ-9, we found overlap in symptoms related to suicidal ideation (item 24, “better off dead”) and cognitive problems (items 47 and 48, “difficulty concentrating” and “trouble thinking”). In contrast, PHQ-9 items related to anhedonia, low energy, feelings of failure, and sleep or appetite disturbances were not represented in our scale. This may be due to the overlap of these symptoms with the clinical manifestations of TB, as they can result from the disease itself or its treatment ^24^. Conversely, the prominence of cognitive symptoms (items 47 and 48) among the most discriminative items may indicate their relative specificity in this context, suggesting a potential shift in symptom relevance when screening for depression in individuals undergoing TB treatment.

Compared to previous studies, the scale developed in this study demonstrated superior discriminative performance. Using the CES-D, *Chishinga et al*. reported an auROC of 78% in a TB population, notably lower than the 96% observed in our analysis ^25^. In Turkey, *Aydin et al.* found that a GHQ-12 version yielded sensitivity and specificity of 80.7% and 87.1%, respectively, also lower than the 89.4% and 92.2% reported here ^26^. Among individuals with HIV, *Natamba et al.* reported an auROC of 0.82 using the CES-D ^27^, while *Thai et al.* found an auROC of 0.88 with sensitivity and specificity of 79.8% and 83.0%, respectively, for the optimal cutoff point ^28^.

### Limitations

A key limitation of this study lies in the S-ROC process, which, while aiming to maximize auROC with the fewest items—similar to variable selection in predictive modeling—may compromise domain representativeness. This approach may exclude conceptually relevant items that do not significantly improve ROC performance. The lack of a separate validation sample represents another important limitation, as no equivalent to model calibration was applied, increasing the risk of overfitting to the current sample.

Subgroup analyses were not feasible due to limited sample size, precluding assessment of differential symptom presentation by TB type (e.g., drug-sensitive vs. MDR/XDR) or comorbid conditions such as diabetes or rheumatic diseases, which may influence both depressive symptoms and TB treatment outcomes ^29–31^.

The study population consisted of individuals referred for psychiatric evaluation in Lima Norte, which may have introduced referral bias. Detection bias is possible if some primary care providers lacked adequate training to identify depression, potentially leading to the underrepresentation of individuals with mild or moderate symptoms. To address concerns regarding the length of the 58-item scale, item order was randomized across three versions. The SCID-5 was used as the gold standard to ensure standardized diagnostic criteria, although clinical judgment could have been an alternative reference.

## Data Availability

All data produced in the present study are available upon reasonable request to the authors

## Acknowledgements

The authors thank the PCT personnel in the DIRIS LIMA NORTE, especially those that collaborated with study procedures in the health centers where participants were selected. The second study was partially funded by a CIENCIACTIVA scholarship (National Council for Science, Technology and Technological Innovation) for the Doctoral Program in Epidemiological Research at the School of Public Health and Administration, Universidad Peruana Cayetano Heredia, which supported PRG’s doctoral studies.

## Supplementary Data (Optional)

**Figure S1.**
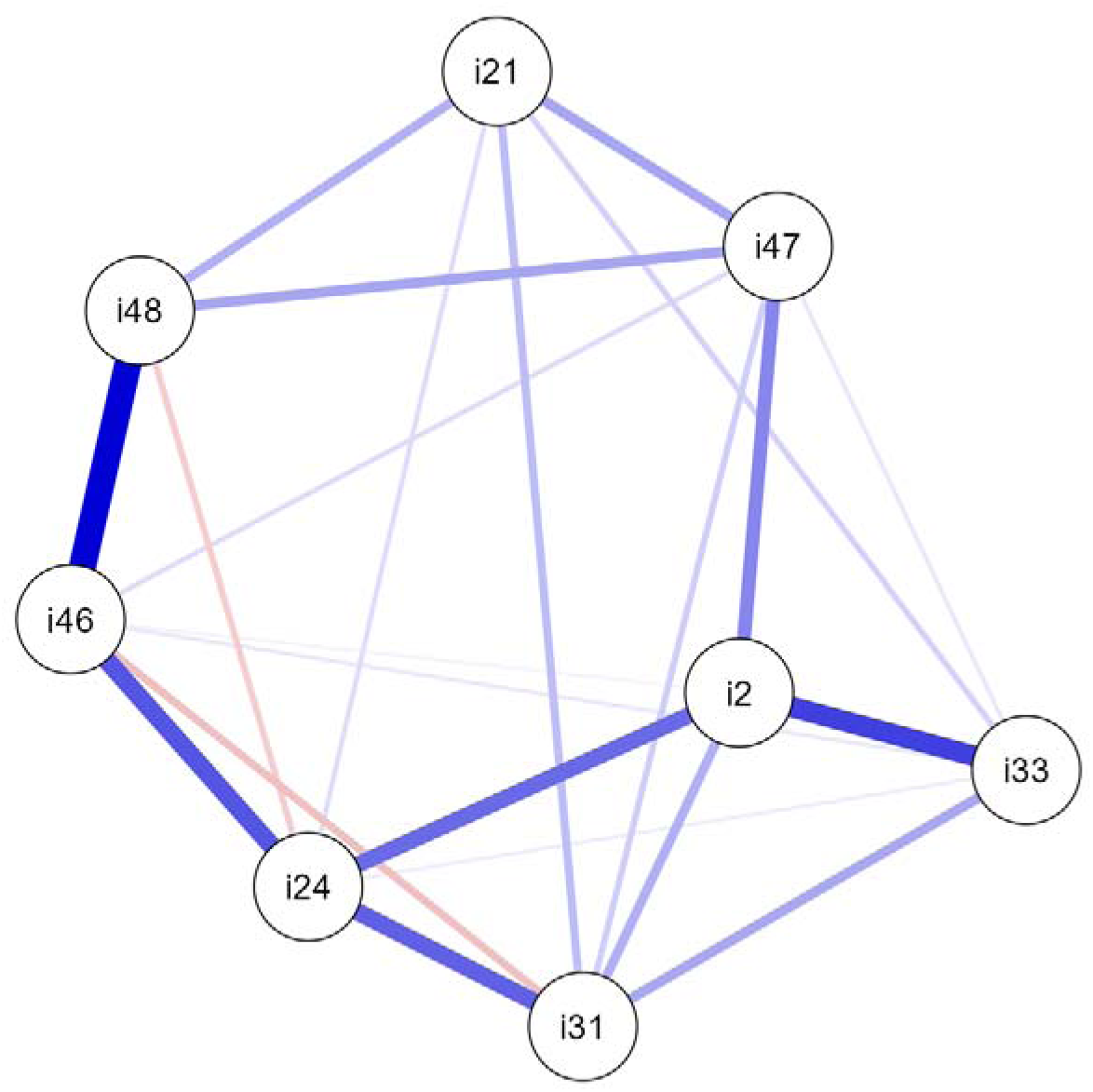
Network Analysis Diagram.

**Additional figures/tables labeled as Table S1, Figure S1, etc.**

## Notes

### Competing Interest Statement

The authors have declared no competing interest.

### Funding Statement

The study was partially funded by a CIENCIACTIVA scholarship (National Council for Science Technology and Technological Innovation) for the Doctoral Program in Epidemiological Research at the School of Public Health and Administration, Universidad Peruana Cayetano Heredia, which supported PRGs doctoral studies.

### Author Declarations

The Institutional Ethics Committee of Universidad Peruana Cayetano Heredia from Lima, Peru approved the research protocol by the Constance number 683 - 36 - 20, dated April 5th 2021

