## Supplementary figures and images for "Validation and Psychometric Properties of a Depression Scale for People with Tuberculosis in Lima, Peru"

### Supplemental Figure 1

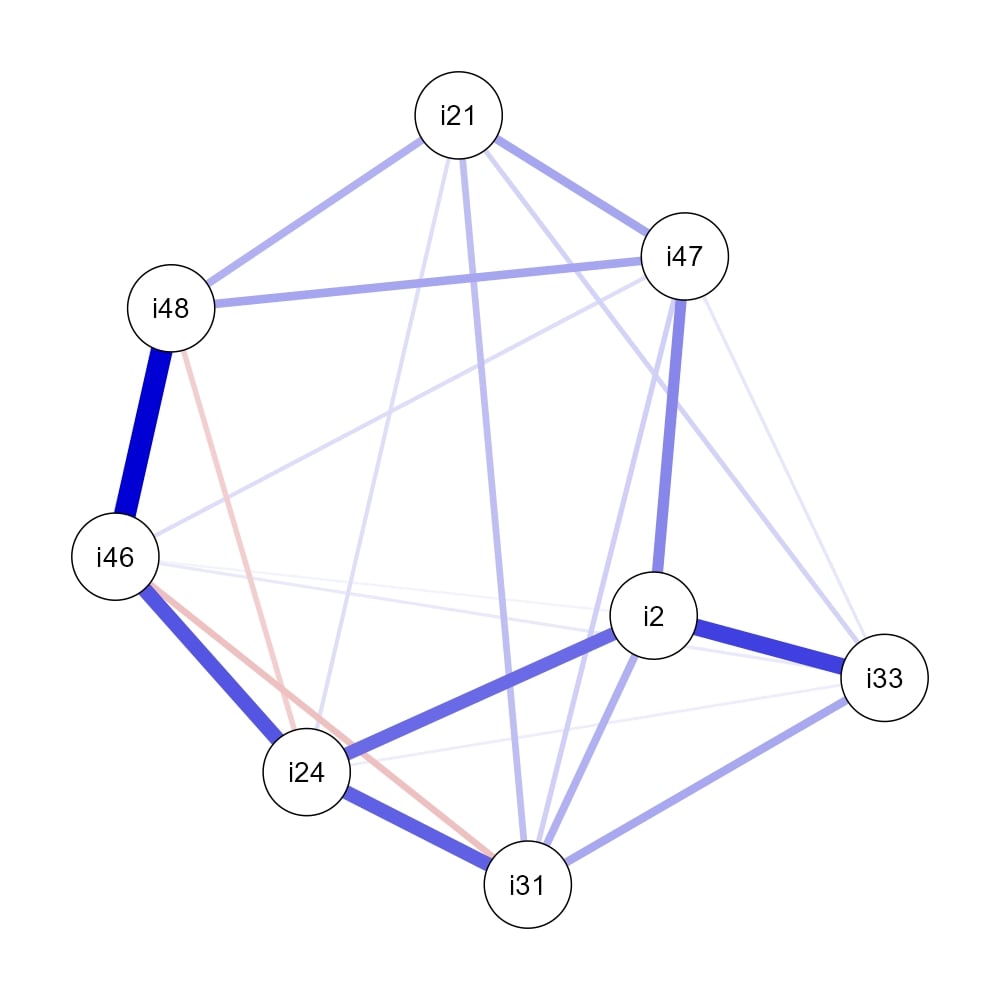
